# Therapy-Associated Oncogenic Axes of Core Cellular and Metabolic Programs Revealed by Multi-Omics Regulatory Network Analysis in Adrenocortical Carcinoma

**DOI:** 10.64898/2025.12.21.25342239

**Authors:** Javad Omidi

## Abstract

Adrenocortical carcinoma (ACC) is a rare and aggressive malignancy with poor prognosis and limited therapies. To elucidate its post-transcriptional regulation, ceRNA networks were reconstructed from TCGA-ACC and GTEx 2025 data. Tumor networks exhibited compact topology and tumor-specific hub miRNAs (miR-466, miR-940, miR-507), which were downregulated, inversely correlated with oncogenic transcripts, and associated with adverse survival. Candidate targets were identified through dual validation by miRTarBase and miRDB, further confirmed with TargetScan, and filtered for inverse correlation, significant upregulation in tumors, and treatment-response association. The intersection of these stringent layers yielded four robust oncogenic biomarkers; CKS2, ERP44, ERG28, and FAM32A that were consistently upregulated and annotated to key processes: chromatin remodeling and cell cycle control (CKS2), apoptosis regulation (FAM32A), ER proteostasis and redox balance (ERP44), and sterol biosynthesis (ERG28). Network analysis highlighted CKS2 as central PPI hubs. Prognostic modeling demonstrated strong survival divergence, with CKS2 conferring the highest risk (HR = 4.49, p = 1.1e-06), while integration of the four-gene panel achieved near-perfect classification accuracy (AUC = 0.99). Collectively, these findings define CKS2, ERP44, ERG28, and FAM32A as high-confidence biomarkers in ACC, derived through multi-layered overlap filtering, and underscore their diagnostic and prognostic relevance.

## 1. Introduction

Adrenocortical carcinoma (ACC) is a rare endocrine malignancy that originates from the adrenal cortex, characterized by aggressive clinical behavior and poor prognosis [1, 2]. With an estimated annual incidence of 0.7-2 cases per million individuals, ACC accounts for only a minor proportion of adrenal tumors but is responsible for the majority of adrenal cancer-related deaths due to its high metastatic capacity and resistance to conventional therapies [2]. Despite significant improvements in surgical techniques and the adoption of adjuvant therapeutic modalities, the five-year survival rate for advanced-stage disease remains below 40% [3, 4]. This poor outcome largely reflects delayed diagnosis, frequent recurrence, and the limited effectiveness of currently available systemic therapies. These limitations have created a strong impetus for identifying robust molecular biomarkers that can enhance early detection, allow accurate risk stratification, and guide personalized therapeutic decision-making in ACC [5].

The central role of biomarkers in enabling personalized medicine and accelerating drug development has been extensively emphasized in recent reviews. McBrearty et al. underscored that companion diagnostics based on robust biomarkers are indispensable for patient stratification, therapeutic decision-making, and the successful implementation of precision oncology [6].Recent years have witnessed a paradigm shift in cancer biology, with non-coding RNAs (ncRNAs) increasingly recognized as pivotal regulators of gene expression and tumorigenesis [7]. Within this diverse class, microRNAs (miRNAs) and long non-coding RNAs (lncRNAs) have attracted substantial attention due to their ability to modulate both oncogenic signaling and tumor suppressor pathways across a wide spectrum of malignancies, including ACC [5, 8, 9].

The competing endogenous RNA (ceRNA) hypothesis provides a mechanistic explanation for these interactions, proposing that transcripts harboring shared miRNA response elements can influence each other’s expression by competitively binding common miRNAs. Through this mechanism, ceRNA networks form highly interconnected layers of post-transcriptional regulation [10]. Such regulatory crosstalk has been shown to drive fundamental cancer hallmarks, including uncontrolled proliferation, metastatic dissemination, and resistance to therapeutic agents, thereby establishing a conceptual foundation for the identification of novel biomarkers and the development of targeted therapeutic strategies [11]. Similar integrative approaches in other cancers have revealed miRNA-mRNA regulatory axes with strong clinical implications. For example, Duan et al. demonstrated that miR-942-5p-mediated downregulation of PIEZO1 promotes NSCLC progression through MAPK signaling, highlighting the potential of ceRNA interactions in shaping tumor prognosis [12].

Systems biology approaches, particularly network-based analyses, have transformed cancer research by allowing the integration of diverse multi-omics datasets and providing insights into the complexity of gene regulation at a systems-wide level [13]. Through the construction and interrogation of ceRNA networks, it becomes possible to identify central nodes, genes or RNAs with high connectivity or regulatory influence that may function as master regulators or clinically relevant biomarkers [14]. Such network-based strategies have already proven effective in other malignancies, where ceRNA analyses have identified lncRNAs and mRNAs with diagnostic and prognostic potential in breast, lung, and colorectal cancers [15]. Nevertheless, despite the promise of this methodology, comprehensive ceRNA network investigations in ACC are still limited, and the regulatory significance of individual genes within these networks has not yet been systematically delineated [5].

Recent ceRNA-based studies have highlighted the utility of systems-level regulatory modeling for biomarker discovery in ACC. Omidi identified synergistic oncogenic ceRNA modules centered on CKS2 and ACAT2, linking cell-cycle progression and metabolic reprogramming with strong diagnostic and prognostic value [16], and uncovered miR-466, miR-507, and miR-665 as central miRNA hubs driving tumor-specific regulatory rewiring, with tumor-suppressive potential [17, 18]. Furthermore, multi-omics and network-based biomarker discoveries have been synthesized in an integrative review, outlining key translational gaps in ACC [19].

Several recent investigations have begun to shed light on the molecular landscape of ACC. The Cancer Genome Atlas (TCGA) [20] has provided a comprehensive transcriptomic and genomic characterization, highlighting recurrent alterations in canonical oncogenic pathways such as Wnt/β-catenin and p53/Rb [5, 21]. Parallel studies focusing on non-coding RNAs have reported aberrant expression of specific miRNAs, with miR-483-5p and miR-195 emerging as potential diagnostic and prognostic biomarkers in ACC [22, 23].

Likewise, lncRNAs including HOTAIR and MALAT1 have been implicated in tumor progression, metastatic dissemination, and adverse clinical outcomes [5]. Despite these important advances, the integration of such findings into a unified systems-level ceRNA framework, capable of uncovering novel regulatory hubs and clinically relevant biomarkers in ACC remains largely underexplored.

Given this critical gap, the present study presents a completely novel, integrative framework for biomarker discovery in ACC, achieved through the construction and analysis of a ceRNA network based on mRNA and miRNA expression profiles from newly released publicly available datasets TCGA-ACC [20] and GTEx 2025 [24]. Importantly, all identified regulatory interactions and candidate biomarkers uncovered here are reported for the first time in ACC and represent previously uncharacterized features of its molecular landscape. Our approach uniquely combines experimentally validated interaction databases with network centrality analyses specifically tailored for ACC, enabling the systematic discovery of key molecular signatures for diagnosis, prognosis, and therapeutic targeting. To our knowledge, no prior study has generated or characterized these interactions in ACC using this comprehensive systems-level methodology. These findings thus substantially enrich the molecular toolkit available for ACC management and provide a strong foundation for future functional and clinical validation in this rare and aggressive malignancy.

## 2. Materials and Methods

### 2.1 Data Acquisition and Preprocessing

Transcriptomic profiles, including both gene and microRNA expression, were collected from The Cancer Genome Atlas (TCGA) [20] for 78 ACC tumor samples and from the Genotype-Tissue Expression (GTEx 2025) project [24] for 250 normal adrenal tissues. To maintain consistency between datasets, potential batch effects were examined, and correction was carried out using the `removeBatchEffect` function of the limma package [25], which applies linear modeling to adjust for batch-related variation while preserving true biological signals.

### 2.2 Quality Control and Filtering Strategies

Data integrity and uniform expression across samples were ensured by subjecting both mRNA and miRNA datasets to a multi-step quality control and filtering process. Genes with Transcripts Per Million (TPM) and miRNAs with Reads Per Million (RPM) values less than 1 in more than 80% of samples were excluded. Sequencing biases were further minimized through normalization using the `TMM` method, followed by `voom` log-transformation. Post-normalization evaluations confirmed that no major deviations or technical artifacts were present. All filtering procedures were implemented with custom R scripts, maintaining consistent thresholds across datasets and establishing a solid foundation for subsequent integrative and network analyses.

Because of the limited sample size in TCGA-ACC [20], relatively flexible thresholds were applied in order to minimize technical noise while still retaining rare transcripts with potential regulatory importance, such as miRNAs capable of exerting strong regulatory effects [26, 27]. To further control residual noise, downstream procedures included normalization, batch effect correction, and multi-step validation encompassing cross-dataset expression consistency, ceRNA network context, and survival analysis. Through these steps, the retained features were ensured to represent robust and biologically meaningful signals.

### 2.3 Differential Expression Analysis

Differential expression analysis between tumor and normal adrenal tissue samples was carried out with the widely used `limma-voom` framework [25], which combines linear modeling and empirical Bayes moderation after estimating mean-variance relationships by applying precision weights to log-transformed counts through the `voom` method. This approach is broadly acknowledged as one of the most reliable strategies for RNA-Seq transcriptomic profiling. During preprocessing, raw RNA-Seq counts from tumor (TCGA-ACC [20]) and normal (GTEx 2025 [24]) cohorts were filtered to retain only protein-coding genes, and transcript version suffixes were removed. The datasets were subsequently aligned using Ensembl gene identifiers, and unstranded count data were merged into a single expression matrix across all samples for downstream analyses. To enhance model stability and reduce noise, lowly expressed genes were filtered through the `filterByExpr` function, which incorporates the experimental design into filtering. Library size normalization was then performed with the `TMM` (trimmed mean of M-values) method to correct for compositional biases before applying linear modeling.

A design matrix was generated to represent the two experimental groups (normal vs. tumor), with normal samples designated as the reference category. Linear modeling was performed using the `lmFit` function in the `limma` package, and variance estimates were stabilized through empirical Bayes moderation implemented by the `eBayes` procedure. Differentially Expressed Genes (DEGs) were determined with the `topTable` function, where significance was ranked according to Benjamini-Hochberg adjusted p-values. Genes were classified as differentially expressed when meeting the criteria of an adjusted p-value < 0.05 together with an absolute log2 fold change greater than 1. After correction and standardization of gene identifiers, the final DEG list was exported to ensure compatibility with subsequent downstream analyses.

### 2.4 Integration of miRNA-mRNA Interactions and ceRNA Network Construction

A ceRNA network was comprehensively constructed using transcriptomic data from TCGA-ACC tumor samples and GTEx normal adrenal tissues. Experimentally validated miRNA-mRNA interactions were obtained through the `multiMiR` R package [28], which integrates high-confidence interactions from databases including miRTarBase, TarBase, and miRecords. Interactions supported by at least five distinct sources were retained to maximize reliability.

For adrenal-specific functional relevance, each miRNA-mRNA interaction was cross-checked against expression profiles from both tumor and normal tissues, and only those pairs exhibiting significant negative correlation (p < 0.05) were kept. By combining experimental validation with tissue-specific expression data, the biological robustness of the network was reinforced. The final network was restricted to miRNA-mRNA interactions, consistent with current practices in ceRNA research.

### 2.5 Survival Analysis

Overall survival (OS) analysis was performed to assess the prognostic significance of candidate biomarkers in ACC. Clinical information was standardized by extracting case identifiers, follow-up data, and survival outcomes. OS time was defined as the minimum of “days to death” or “days to last follow-up,” expressed in months, with OS event status coded as 1 (death) or 0 (censored). For each miRNA, expression values were log2-transformed and separated into high- and low-expression categories using the median as the cutoff. Kaplan-Meier survival curves were generated with the `lifelines` Python package, and group differences were evaluated through log-rank testing. Hazard Ratios (HR) were calculated by comparing event rates between the high- and low-expression groups.

## 3. Results

### 3.1 ceRNA Network Topology and Identification of Hub Genes

The analysis of ceRNA networks derived from normal adrenal and tumor transcriptomes of ACC highlights substantial alterations in regulatory architecture between physiological and cancerous states. The tumor-specific network displays a noticeably more compact and interconnected topology, pointing to extensive regulatory rewiring in ACC. In this network, the main hubs are hsa-miR-466, hsa-miR-940, and hsa-miR-507, none of which serve as central or even active components under normal conditions. The presence of these tumor-exclusive hubs signifies a marked shift in the balance and influence of miRNA-mediated regulation under oncogenic pressure [17, 18].

These observations are further substantiated by quantitative analysis of network centrality, revealing a pronounced shift in hub miRNAs between normal and tumor states. In the tumor network, miR-466 emerges as the dominant hub with the highest betweenness centrality, followed by miR-940 and miR-507, all of which show negligible or no centrality in the normal network [17, 18]. These reciprocal changes highlight a pronounced rewiring of regulatory interactions in ACC, suggesting that miR-466, miR-940, and miR-507 may gain tumor-specific regulatory influence. The miRNAs identified as central hubs in ACC ceRNA networks have also been implicated in the regulatory landscapes of various other cancers [29–32].

### 3.2 Functional Connectivity Among Hub miRNAs and Target Genes Discovery

Independent transcriptomic and survival analyses of ACC datasets have identified miR-466, miR-940, and miR-507 as potential tumor-suppressive miRNAs in this malignancy [17, 19]. Based on the oncogenic transcripts they target, their pathway involvement, expression patterns across clinical stages, and Kaplan-Meier survival analyses, these candidates were identified as tumor suppressors collectively, representing the integrated outcome of multiple lines of evidence.

miR-466 exhibits tumor-suppressive effects in colorectal cancer (CRC) by inhibiting proliferation, migration, and invasion through downregulation of cyclin D1 and MMP-2, while promoting apoptosis via BAX upregulation, with low expression correlating with advanced Tumor Node Metastasis (TNM) stages and poor prognosis [29]. In prostate cancer, miR-466 suppresses bone metastasis by directly targeting RUNX2, impairing oncogenic functions and improving survival outcomes [30]. hsa-mir-940 demonstrates context-dependent tumor suppression, showing diagnostic and prognostic value in cancers like gastric cancer and hepatocellular carcinoma by regulating pathways linked to proliferation and metastasis [31]. In gastric carcinoma, miR-507 functions as a tumor suppressor by directly targeting CBX4, thereby inhibiting cell proliferation and invasion, inducing apoptosis, and suppressing the activation of Wnt/β-catenin and HIF-1α pathways [32].

Based on these findings, the regulatory impact of these three miRNAs was further investigated by identifying their putative mRNA targets using two high-confidence resources: miRTarBase [33] and miRDB [34]. All retrieved target lists were then cross-referenced with gene expression data from ACC samples. Target genes were retained only if they showed a significant inverse correlation with their corresponding miRNA (r < -0.3, p < 0.05), reflecting context-specific regulatory interactions in ACC. The resulting filtered gene lists were compiled in **Table 1**, highlighting for each miRNA the most relevant targets in the tumor context. To further prioritize biologically meaningful candidates, genes that were also significantly upregulated in ACC were marked with an asterisk (*), indicating their likely oncogenic function and relevance as downstream effectors of tumor-suppressive miRNA activity.

**Table 1.**
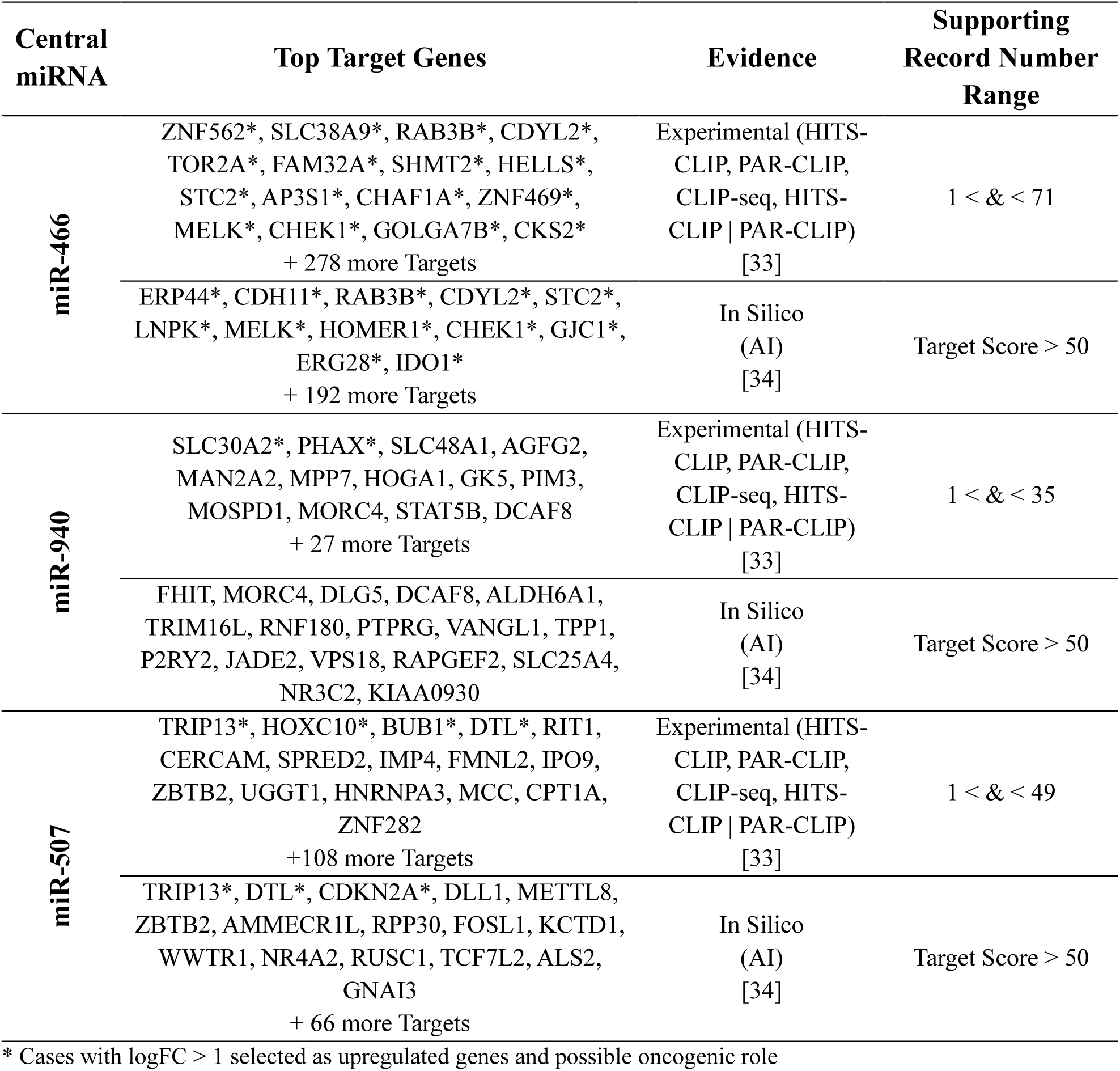
Validated and predicted target genes of tumor-suppressive miRNAs identified in ACC. For each miRNA (miR-466, miR-940, and miR-507), targets were extracted from miRTarBase [33] and miRDB [34]. Genes showing strong meaningful negative correlation with miRNA expression (p < 0.05) in ACC transcriptomes were retained.

The selected target from **Table 1** was further validated by TargetScan [35], yielding 19 potential oncogenes, including RAB3B, FAM32A, MELK, CHEK1, SLC38A9, CKS2, AP3S1, GOLGA7B, ZNF562, CDYL2, STC2, ERG28, GJC1, IDO1, HOMER1, ERP44, TRIP13,

CDKN2A, and DTL. The correlation cutoff of r < -0.3 was chosen to strike a balance between rigor and inclusiveness, as applying a more stringent threshold such as r < -0.5 would eliminate moderately correlated yet biologically meaningful interactions, while a looser criterion could increase the risk of spurious associations. This cutoff, together with p < 0.05, has also been utilized in earlier network-based analyses [36], where the retained miRNA-mRNA pairs typically demonstrated moderate to strong negative correlations within this interval.

### 3.3 Localization of Key Target Genes within the Differential Landscape

To further substantiate the oncogenic potential of the prioritized target genes, their differential expression profiles were visualized using a volcano plot derived from the full transcriptomic comparison between ACC tumor and normal adrenal tissues. The selected target genes after TargetScan filtering, identified as miRNA targets with inverse expression correlation appear among the significantly upregulated genes (log₂FC > 1, p < 0.05). This spatial clustering reinforces their transcriptional activation in ACC and supports the robustness of the dual-filtering strategy employed, which integrates experimental miRNA-mRNA targeting data with expression-based validation.

### 3.4 Protein-Protein Interaction and Functional Enrichment of Upregulated Key Target Genes

The protein-protein interaction (PPI) network of the prioritized target genes was constructed using the STRING database [37] with a medium confidence threshold of 0.4. Disconnected nodes were excluded to ensure the biological relevance of the network, and the visualization was restricted to a maximum of five interactors in the first shell to minimize noise from indirect associations while retaining the most informative connections. This approach was applied to enhance interpretability by focusing on the most biologically meaningful interactions. The final network contained 24 nodes and 48 edges, with an average node degree of 4 and an average local clustering coefficient of 0.4. Compared to the expected number of edges (10), the observed enrichment was markedly higher, yielding a highly significant PPI enrichment p-value of < 1.0e-16, indicating that the selected genes are functionally connected beyond random chance (**Figure 1a**).

**Figure 1.**
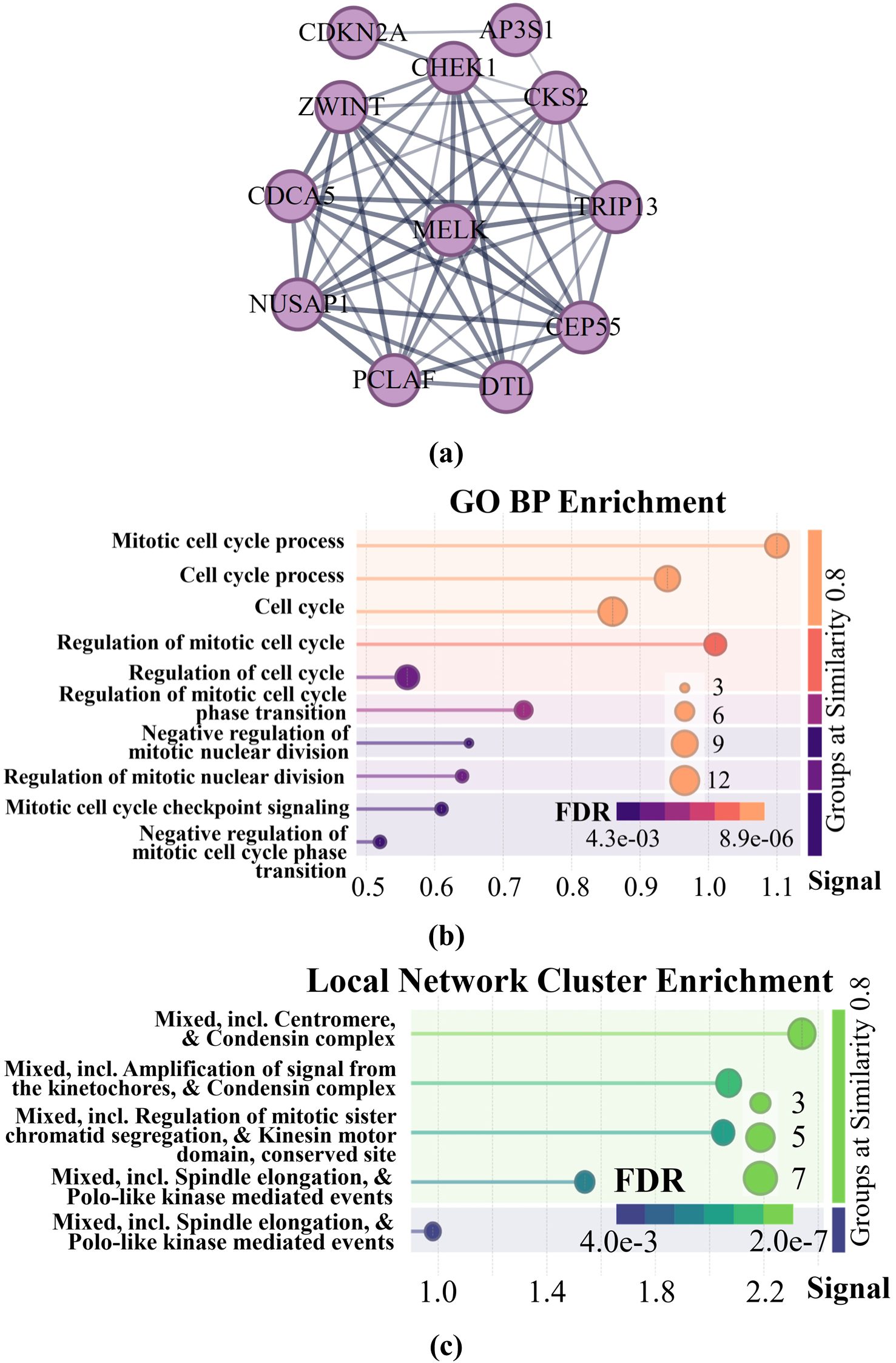
Enrichment Analysis on selected oncogenes (a) PPI network of the key target genes using STRING [37]. (b) GO biological process enrichment analysis of the key target genes. (c) Local network cluster enrichment analysis of the key target genes.

Subsequent network topology analysis highlighted key hub genes. Degree centrality identified CKS2 and CHEK1 (degree = 10), followed by TRIP13, DTL, and MELK (degree = 9) as the most connected nodes, underscoring their extensive interaction capacity. Betweenness centrality revealed CKS2 and CHEK1 (0.1545 each) as critical bottlenecks controlling information flow, whereas AP3S1 and CDKN2A (0.0091 each) appeared as secondary mediators. Closeness centrality further emphasized CKS2 and CHEK1 (0.9167 each) as the most proximally located within the network, followed closely by TRIP13, DTL, and MELK (0.8462 each), reflecting their roles as central integrators of molecular interactions. These results indicate that CKS2 and CHEK1 emerge as dominant hubs with strong oncogenic potential, while TRIP13, DTL, and MELK also occupy critical positions within the network architecture. The combination of high connectivity and centrality metrics supports the hypothesis that these genes function as pivotal drivers in tumor-associated molecular processes.

**Figure 1b** illustrates the top ten enriched Gene Ontology (GO) biological processes associated with the prioritized target genes, highlighting the predominance of cell cycle-related terms. The most significantly enriched categories included cell cycle (GO:0007049), mitotic cell cycle process (GO:1903047), cell cycle process (GO:0022402), and regulation of mitotic cell cycle (GO:0007346), all of which underscore the pivotal role of these genes in maintaining proliferative capacity. Additional terms such as regulation of mitotic cell cycle phase transition (GO:1901990), mitotic cell cycle checkpoint signaling (GO:0007093), and negative regulation of mitotic nuclear division (GO:0045839) suggest that these genes are deeply involved in the control of mitotic fidelity and checkpoint integrity, processes whose disruption is strongly linked to genomic instability and oncogenic transformation. The functional annotation further emphasized categories like cell division (GO:0051301), nuclear division (GO:0000280), and positive regulation of cell cycle (GO:0045787), all pointing toward aberrant proliferative signaling as a central mechanism. Given the highly aggressive nature of ACC, enrichment of processes tied to unchecked cell cycle progression and defective checkpoint regulation strongly supports the oncogenic potential of the identified genes. This suggests that the deregulation of mitotic control, coupled with altered stress response pathways, may represent key molecular drivers contributing to tumor initiation and progression in ACC.

The STRING local network cluster enrichment analysis highlights groups of functionally related modules formed by the prioritized genes, as illustrated in **Figure 1c**. The top enriched clusters were associated with the centromere and condensin complex, amplification of kinetochore-derived signals, and regulation of mitotic sister chromatid segregation, as well as spindle elongation and Polo-like kinase-mediated events. These clusters emphasize the strong involvement of the candidate oncogenes in the control of chromosome segregation, spindle dynamics, and mitotic progression. Dysregulation of such processes is widely recognized as a driver of chromosomal instability and aneuploidy, both of which contribute to oncogenic transformation. In the context of ACC, the enrichment of condensin- and kinetochore-associated modules underscores a mechanistic link between aberrant mitotic regulation and the aggressive phenotype of the disease, reinforcing the oncogenic potential of the identified targets.

### 3.5 Treatment-Associated Expression of Key Target Genes

To evaluate the clinical relevance of the selected target genes from **Table 1**, normalized expression levels were visualized across treatment subgroups using clinical metadata from the TCGA-ACC [20] cohort. Specifically, heatmaps were generated to illustrate gene expression patterns in patients who received either pharmaceutical therapy alone or combined pharmaceutical and radiation therapy. As shown in **Figure 2a**, gene expression values were independently min-max normalized within each treatment group to ensure comparability. The resulting heatmaps revealed distinct expression signatures across subgroups. Notably, CKS2, FAM32A, ERG28, and ERP44 displayed consistently elevated expression in both therapy groups, underscoring their potential role as treatment-responsive oncogenic regulators. In the analysis pipeline, patients were stratified into two clinical subgroups: (i) those who had received both radiation and pharmaceutical therapy, and (ii) those who had received pharmaceutical therapy only. For each candidate gene, expression values were compared between the two groups, and the log fold-change (logFC), p-value (two-sided Welch’s t-test), and an integrated EffectScore (calculated as logFC × -log10 p-value) were derived. Genes showing strong differential expression between treatment groups were designated as ‘treatment-response-related’ during the screening step.

**Figure 2.**
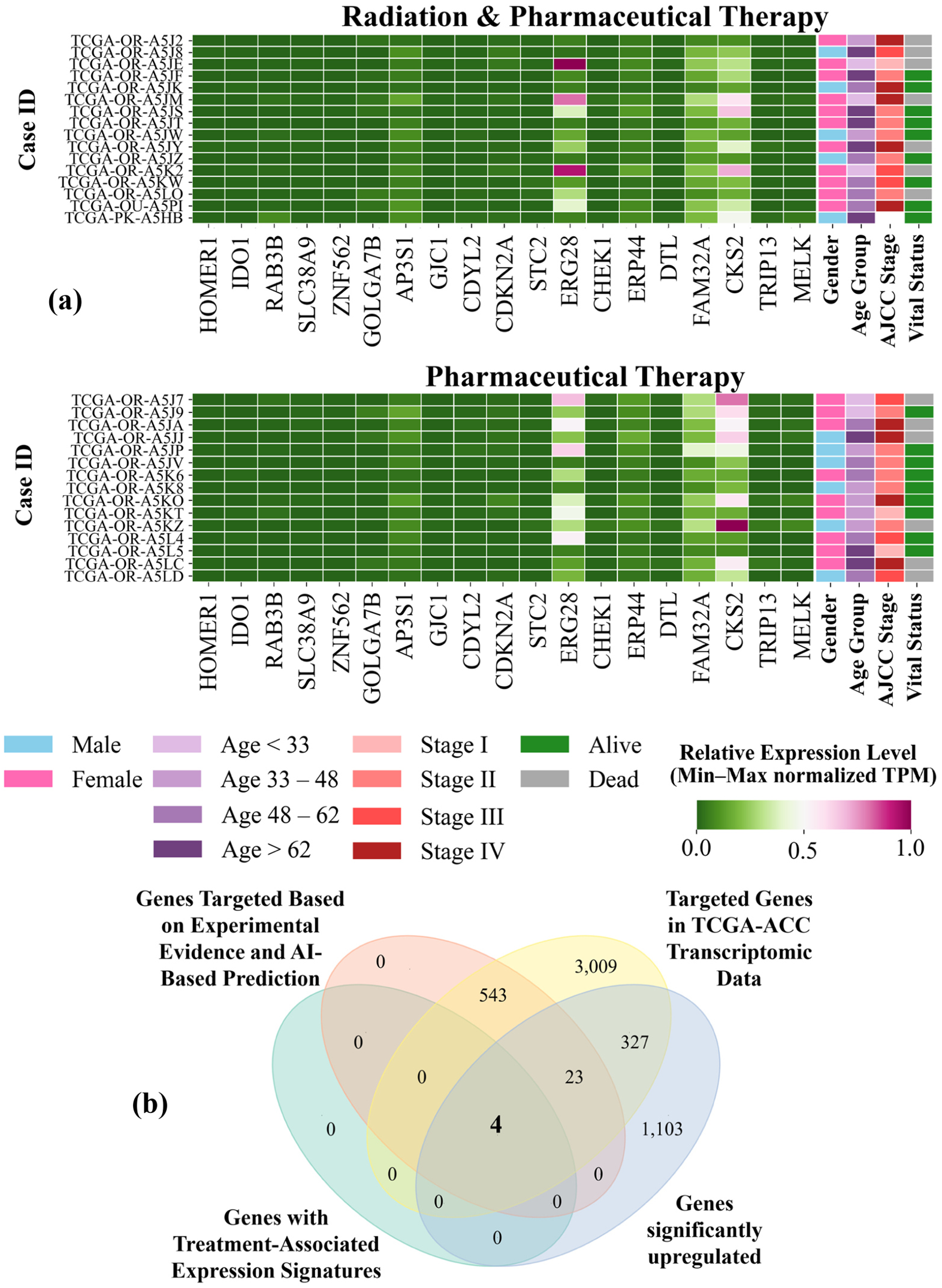
(a) Heatmaps illustrating the normalized expression of selected target genes across ACC patients receiving different therapies. Top: patients undergoing both radiation and pharmaceutical therapy; bottom: patients treated exclusively with pharmaceutical therapy. Color intensity reflects min-max normalized expression levels across genes within each group. Clinical annotations for gender, age group, AJCC stage, and vital status are displayed alongside the expression matrices. (b) Venn diagram illustrating the integrative filtering process used to identify final oncogenic candidates. Four genes were prioritized based on overlap between experimental/computational-based targeting, TCGA-ACC [20] transcriptomic validation, significant upregulation, and association with clinical response to therapies.

In addition to gene expression profiles, **Figure 2a** integrates clinical features; including gender, age categories (< 33, 33 - 48, 48 - 62, > 62 years), AJCC stage, and vital status; allowing a stratified view of therapy response across patient subgroups. Within the radiation plus pharmaceutical therapy group, female patients exhibited generally higher expression of CKS2, FAM32A, ERP44, and ERG28, particularly in Stage II cases that remained alive (e.g., TCGA-OR-A5JS, TCGA-OR-A5JT). Male patients in this group tended to show lower expression intensities, with survivors often restricted to Stage II or undefined cases (e.g., TCGA-OR-A5JZ, TCGA-PK-A5HB). Age-stratified inspection suggests that patients younger than 48 years, particularly women, more frequently demonstrated elevated expression of treatment-responsive genes and favorable survival, whereas older patients (>62 years) in advanced stages exhibited poorer outcomes. In contrast, the pharmaceutical therapy-only group displayed more consistent upregulation of the four genes across both sexes, yet female patients again showed stronger expression signals coupled with better survival, particularly in Stage I-II disease (e.g., TCGA-OR-A5J9, TCGA-OR-A5KT, TCGA-OR-A5L5). Notably, male patients in Stage IV or late Stage III frequently exhibited high expression but poor prognosis (e.g., TCGA-OR-A5JJ, TCGA-OR-A5LD), underscoring a potential sex-specific disparity in therapeutic benefit. These patterns indicate that younger females with earlier-stage tumors derived the most apparent benefit, both in terms of gene activation and survival, whereas older males and those with Stage IV disease showed limited responsiveness, highlighting clinically relevant heterogeneity in treatment outcomes.

In other studies, CKS2 has emerged as a promising therapeutic target in cancer, with studies showing that its overexpression promotes tumor cell proliferation and survival, while silencing or inhibiting CKS2 induces cell cycle arrest, apoptosis, and sensitization to chemotherapy across multiple cancer types, including hepatocellular and gastric cancers [38]. Protein kinase CK2, often overexpressed in tumors, is similarly recognized as a valuable target; its inhibition by agents such as CX-4945 (silmitasertib) has demonstrated anti-tumor activity, both as a monotherapy and in combination with other chemotherapeutics, leading to disease stabilization in clinical trials and ongoing evaluation in diverse malignancies [39–41].

### 3.6 Venn Diagram-Based Gene Filtering

A four-way integrative filtering strategy was applied to prioritize high-confidence oncogenic candidates in ACC:

1. genes targeted based on two experimental evidence and computational-based prediction,
2. transcriptomic validation using TCGA-ACC data [20],
3. significant upregulation in tumor samples,
4. treatment-associated expression of targets with secondary TargetScan validation.

As illustrated in **Figure 2b**, the intersection of these stringent criteria yielded four core genes; CKS2, ERG28, FAM32A, and ERP44, which represent the most robust candidates for further functional and translational investigations. Recent studies have identified CKS2 as a prognostic biomarker in ACC, with elevated expression levels consistently associated with adverse clinical outcomes and reduced overall survival in ACC patients [42]. By contrast, the other three genes (FAM32A, ERP44, and ERG28) remain novel, as their prognostic or functional significance in ACC has not been previously documented.

### 3.7 Expression Profiling of Candidate Oncogenes in ACC

Expression profiling of the four candidate oncogenes; CKS2, ERG28, FAM32A, and ERP44 revealed a clear and statistically significant upregulation in ACC tumor samples compared to normal adrenal tissues, as shown in **Figure 3a**.

**Figure 3.**
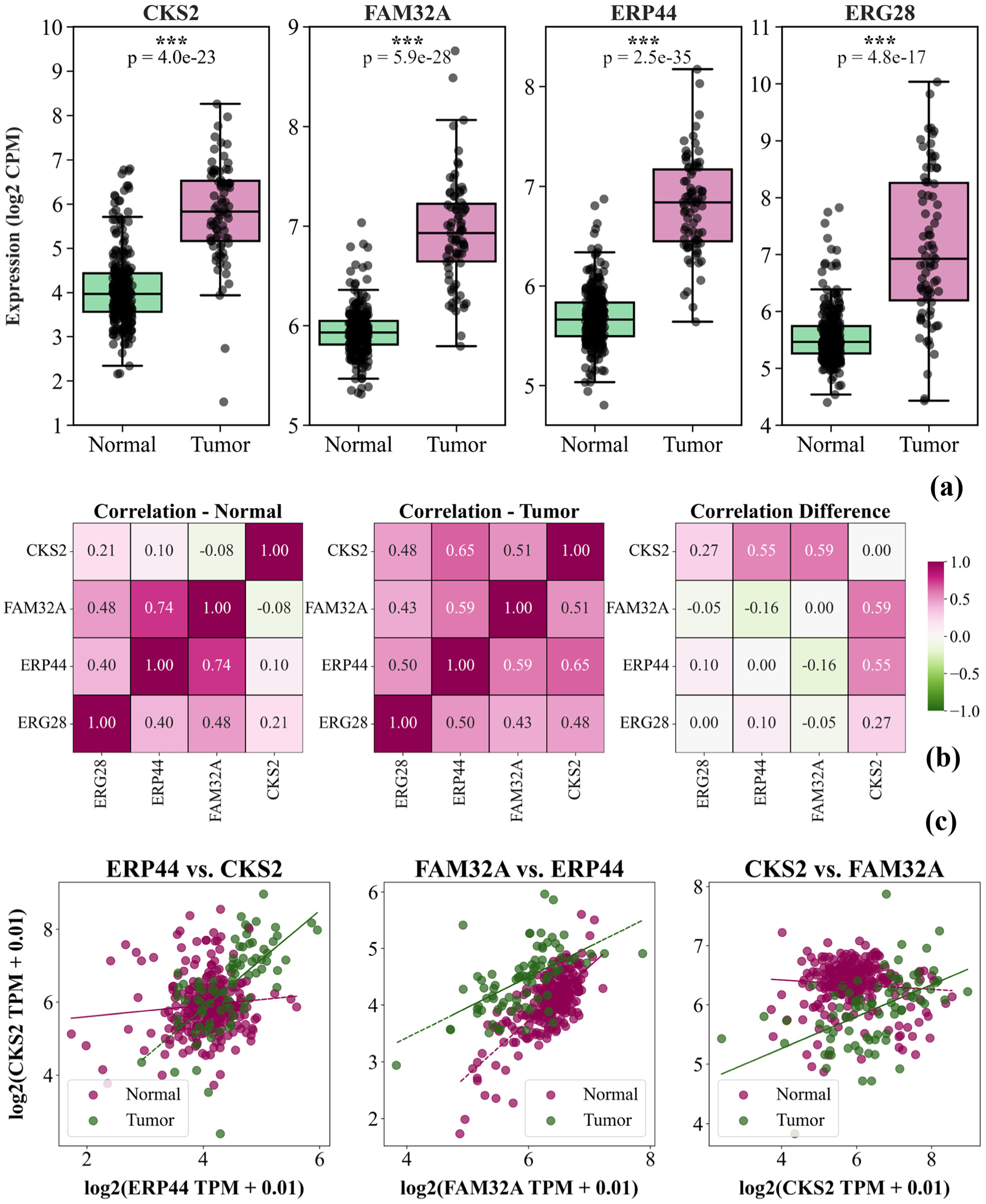
(a) Boxplots showing the expression levels of CKS2, FAM32A, ERP44, and ERG28 in normal versus ACC tumor samples. All four genes exhibit significantly higher expression in tumor tissues (p < 0.001), supporting their potential oncogenic roles in ACC. (b) Condition-specific co-expression of CKS2, FAM32A, ERP44, and ERG28 - Pearson correlation heatmaps in normal (left), tumor (middle), and differential (right) states. (c) Scatter plots of gene pairs colored by condition.

Boxplots demonstrate consistent elevation in transcript levels across all four genes in the tumor group (p < 0.001 for each). Four candidate oncogenes exhibit a compact, unimodal distribution in normal tissues with limited variability, while tumor samples display a marked shift toward higher expression and a pronounced widening of the upper tail suggesting transcriptional activation and heterogeneity among tumor cases.

### 3.8 Condition-Specific Co-regulation Patterns of Candidate Oncogenes

To gain deeper insight into the dynamic regulatory function of the identified genes, the pairwise co-expression patterns of CKS2, ERG28, FAM32A, and ERP44 under normal and tumor conditions were examined. As depicted in **Figure 3b**, notable differences in gene-gene associations between the two states are revealed by the Pearson correlation matrices. In the correlation heatmaps (**Figure 3b**), distinct rewiring of co-expression patterns among the candidate oncogenic biomarkers was observed between normal and tumor tissues. In normal samples, ERP44 and FAM32A displayed a strong positive correlation (r ≈ 0.74), whereas CKS2 exhibited weak or negative associations with these genes (r ≈ 0.10 with ERP44 and r ≈ -0.08 with FAM32A). By contrast, in tumor tissues, CKS2 gained substantial connectivity, showing moderate to strong positive correlations with both ERP44 (r ≈ 0.65; Δr ≈ +0.55) and FAM32A (r ≈ 0.51; Δr ≈ +0.59), while the ERP44-FAM32A interaction was slightly attenuated (Δr ≈ -0.16). ERG28 remained positively correlated with all partners in both states, although its changes were comparatively modest. To highlight these tumor-specific shifts, scatter plots were generated for the pairs with the greatest differential correlations (**Figure 3c**), illustrating that the most prominent gains in association involved CKS2 with ERP44 and FAM32A, thereby underscoring the emergence of CKS2-centered transcriptional modules in the malignant context.

### 3.9 Survival Analysis of Candidate Oncogenes

To assess whether the identified genes exert prognostic relevance in ACC, Kaplan-Meier survival analysis was performed based on expression levels of CKS2, ERG28, FAM32A, and ERP44. Patients were stratified into high- and low-expression groups using median cutoffs, and survival differences were evaluated using log-rank tests (**Figure 4a**). In all cases, patients with high expression levels exhibited markedly reduced overall survival compared with those in the low-expression groups.

**Figure 4.**
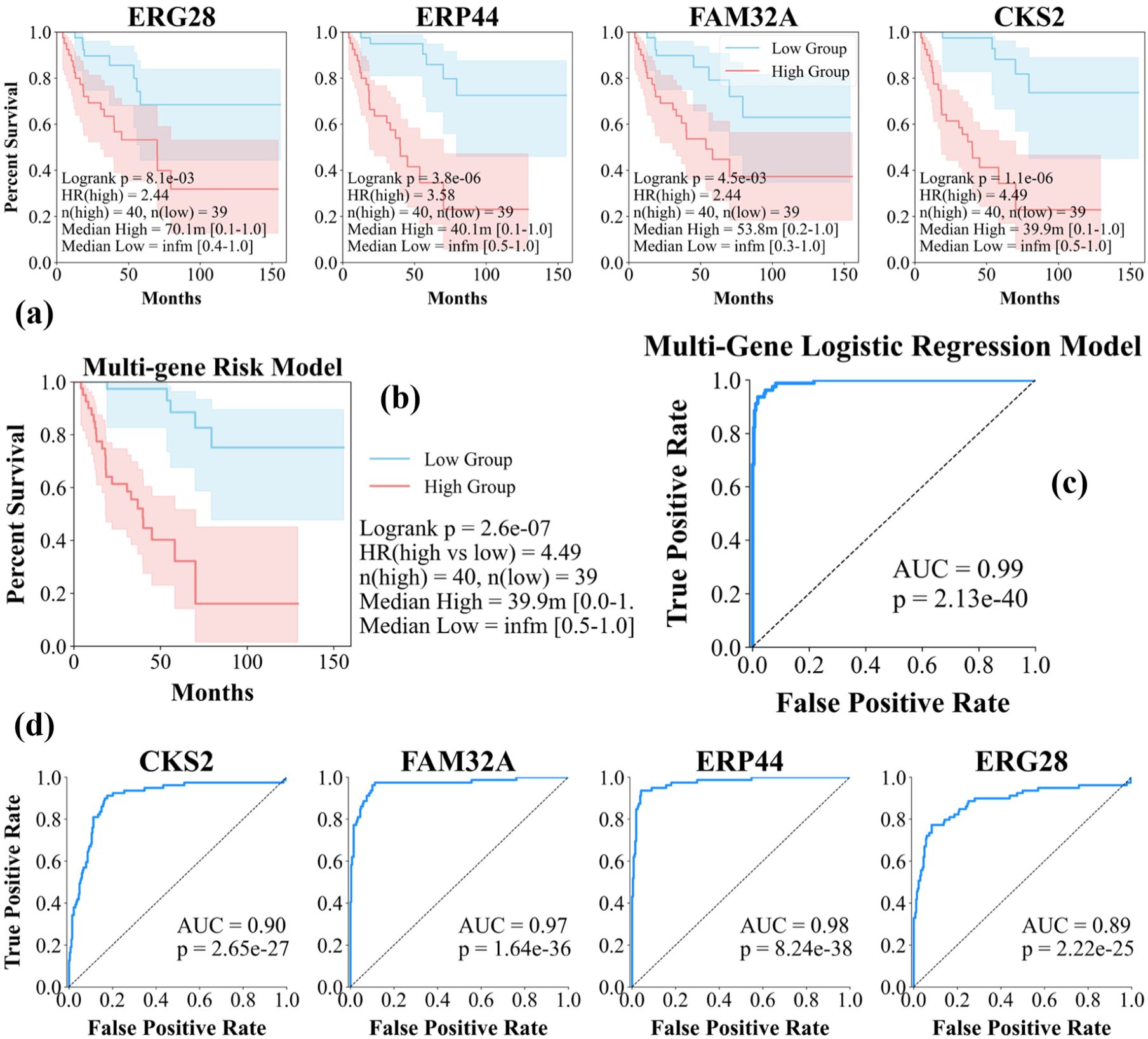
(a) Kaplan-Meier survival curves for CKS2, ERG28, FAM32A, and ERP44 in ACC patients. Patients were stratified into high- and low-expression groups based on median expression. (b) Kaplan-Meier survival analysis of the multi-gene risk score showing overall survival differences between high- and low-risk groups. (c) Receiver operating characteristic (ROC) curve of the multi-gene logistic regression model. (d) ROC curves for CKS2, ERG28, FAM32A, and ERP44 in classifying tumor vs. normal ACC samples.

For ERG28, high expression was associated with significantly poorer survival (logrank p = 8.1e-03), with a hazard ratio (HR) of 2.44. Patients in the high group (n = 40) demonstrated a median survival of 70.1 months, whereas the low group (n = 39) had no definable median survival (infinite), underscoring the detrimental impact of ERG28 upregulation. Similarly, ERP44 high expression correlated with a substantially elevated risk of death (logrank p = 3.8e-06, HR = 3.58). The median survival of the high group was 40.1 months compared with an undefined (infinite) median survival in the low group, reflecting the strong oncogenic potential of ERP44. For FAM32A, overexpression was also significantly linked to worse prognosis (logrank p = 4.5e-03, HR = 2.44). Patients with high expression showed a median survival of 53.8 months, whereas the low group again exhibited no definable median survival, consistent with enhanced tumor aggressiveness in the high-expression cohort. The most pronounced effect was observed for CKS2, where elevated expression conferred a nearly five-fold increased risk of death (logrank p = 1.1e-06, HR = 4.49). The median survival in the high group was only 39.9 months, while the low group displayed no calculable median survival, indicating striking survival divergence. This survival analysis provides compelling evidence that overexpression of ERG28, ERP44, FAM32A, and CKS2 is strongly associated with unfavorable clinical outcomes in ACC. Their consistent predictive power across independent survival metrics highlights their potential oncogenic roles and positions them as promising biomarkers for patient stratification and prognostic evaluation.

A multivariate CoxPH model integrating the four oncogenes was applied, and patients were dichotomized by median risk score. Kaplan-Meier analysis revealed significant survival divergence between high- and low-risk groups (**Figure 4b**). The analysis demonstrated a highly significant prognostic separation between the groups (logrank p = 2.6e-07). Patients classified into the high-risk group (n = 40) exhibited a median overall survival of 39.9 months, whereas the low-risk group (n = 39) showed no definable median survival (infinite), reflecting prolonged survival beyond the maximum follow-up time. The hazard ratio indicated that patients in the high-risk group had a 4.49-fold greater risk of death compared with those in the low-risk group. The markedly divergent survival outcomes underscore the synergistic prognostic value of these genes when considered in combination, beyond their individual contributions. Notably, the multi-gene Cox model showed an exceptionally strong correlation with CKS2 expression (ρ = 0.97), suggesting that the prognostic power of the composite risk score was predominantly driven by CKS2, which emerged as the principal determinant within this gene set. To further evaluate the independent contribution of ERG28, ERP44, and FAM32A, the multivariate model was re-estimated without CKS2, which still yielded a significant separation of survival outcomes (logrank p = 3.8e-06; HR = 3.58). The integrated expression of ERG28, ERP44, FAM32A, and CKS2 defines a robust risk model with strong potential to stratify patients clinically and to highlight their oncogenic relevance in ACC.

### 3.10 Predictive Power Assessment via ROC Analysis

To enhance discriminatory performance, all four candidate oncogenes (CKS2, FAM32A, ERP44, and ERG28) were jointly evaluated through a multivariate logistic regression framework, in which standardized expression values were integrated to derive patient-specific risk scores (**Figure 4c**). The model’s predictive capacity was quantified via ROC curve analysis, revealing near-perfect accuracy with an AUC of 0.99 and highly significant separation between tumor and normal cohorts (p = 2.13e-40).

As illustrated in **Figure 4c**, the multi-gene integration markedly outperformed individual gene assessments, underscoring the synergistic diagnostic value of this composite model. In **Figure 4d**, ROC curve analysis demonstrated strong discriminatory capacity for all four candidate oncogenes. ERP44 achieved the highest diagnostic accuracy with an AUC of 0.98 (p = 8.24e-38), closely followed by FAM32A with an AUC of 0.97 (p = 1.64e-36). CKS2 also exhibited robust predictive performance with an AUC of 0.90 (p = 2.65e-27), while ERG28 showed slightly lower but still substantial accuracy (AUC = 0.89, p = 2.22e-25).

### 3.11 Functional and Pathway-Level Annotation of Candidate Biomarkers

Beyond differential expression analysis, functional and pathway-level annotation of the candidate oncogenes CKS2, FAM32A, ERP44, and ERG28 were conducted to elucidate their mechanistic relevance. Gene Ontology (GO) terms covering all three categories; Biological Process (BP), Cellular Component (CC), and Molecular Function (MF) were retrieved from UniProt [43], providing comprehensive insight into each gene’s functional roles, subcellular localization, and molecular activity.

The GO annotations associated with the prioritized oncogenes further highlighted their potential contributions to tumorigenesis in ACC. CKS2 was found to be enriched for functions related to chromatin binding, histone binding, protein kinase binding, ubiquitin binding, and cyclin-dependent kinase activation, alongside biological processes such as cell division, fibroblast proliferation, meiosis I, mitotic cell cycle phase transition, and regulation of transcription by RNA polymerase II. These annotations indicate a central role for CKS2 in chromatin remodeling, transcriptional regulation, and cell cycle progression, processes whose deregulation is classically associated with uncontrolled proliferation in cancer.

For FAM32A, molecular annotation revealed RNA binding capacity, while its biological process involvement was linked to the apoptotic pathway. Such a profile suggests that FAM32A may exert dual effects on gene expression regulation and cell fate determination. The association with apoptosis is of particular relevance, as perturbations in this pathway may facilitate apoptotic evasion, a hallmark of oncogenic transformation. The annotation landscape of ERP44 emphasized functions in protein disulfide isomerase activity and processes including cell redox homeostasis, glycoprotein metabolism, protein folding, and responses to endoplasmic reticulum (ER) stress and unfolded protein accumulation. These findings implicate ERP44 in proteostasis and redox balance, both of which are frequently hijacked by cancer cells to maintain viability under conditions of rapid growth and metabolic stress. Such activities may provide a selective advantage to malignant cells within the adrenal microenvironment.

Finally, ERG28 was annotated with functions in identical protein binding and protein-macromolecule adaptor activity and was linked to the sterol biosynthetic process. Since sterol metabolism and membrane biogenesis are essential to sustain high proliferative rates, dysregulation of this pathway may contribute to the metabolic reprogramming observed in aggressive ACC tumors. These candidate oncogenes converge on pathways central to malignant transformation, including cell cycle control, apoptosis regulation, proteostasis under ER stress, and sterol metabolism. Their functional annotations thereby provide mechanistic plausibility to the hypothesis that CKS2, FAM32A, ERP44, and ERG28 act as oncogenic drivers in ACC and reinforce their potential as both biomarkers and therapeutic targets.

In terms of CC annotations, CKS2 was associated with the cyclin-dependent protein kinase holoenzyme and the SCF ubiquitin ligase complex, ERP44 was localized to the endoplasmic reticulum lumen, and ERG28 was assigned to the endoplasmic reticulum membrane as a multi-pass membrane protein, while no confident CC annotation was available for FAM32A. Moreover, CKS2 is significantly enriched in the Pathways in cancer, consistent with its central role in cell cycle progression and its established contribution to tumorigenesis.

### 3.12 Molecular Features of Candidate Biomarkers in ACC

Cyclin-dependent kinases regulatory subunit 2 (CKS2) encodes a regulatory protein that binds cyclin-dependent kinases (CDKs) and regulates cell cycle progression [43]. The protein participates in CDK-associated complexes and has been localized to the cyclin-dependent kinase holoenzyme and the SCF ubiquitin ligase complex, both of which are central to the regulation of cell cycle progression and proteasomal degradation pathways. Expression analyses indicate that CKS2 is tissue-enhanced in bone marrow and testis, both highly proliferative tissues, suggesting a role in proliferative and endocrine contexts. Dysregulation of CKS2 has been implicated in malignant transformation, including in ACC, a rare but aggressive endocrine malignancy [43]. At the regulatory level, CKS2 is subject to post-transcriptional control by several microRNAs, which underscores its importance in cancer-associated regulatory networks [44].

ERG28 (also known as C14orf1 or NET51) encodes a small multi-pass membrane protein localized to the endoplasmic reticulum, where it participates in sterol and steroid biosynthesis [43]. ERG28 is ubiquitously expressed, with particularly high levels reported in testis and several cancer cell lines [43]. Functionally, its involvement in lipid and steroid metabolic processes supports a potential role in endocrine regulation [44]. Dysregulation of ERG28, reflected in cancer-related overexpression and integration into steroid biosynthetic pathways, may contribute to aberrant metabolic signaling and oncogenesis in endocrine tissues. Within the adrenal context, such alterations could disrupt steroid homeostasis and promote malignant transformation, positioning ERG28 as a candidate oncogenic biomarker in ACC [44].

ERP44 (also known as ER protein 44, TXNDC4, or PDIA10) encodes an endoplasmic reticulum (ER)-resident protein disulfide isomerase involved in protein folding, redox regulation, and ER calcium signaling [43, 45–47]. Its expression is induced under conditions of unfolded protein response (UPR), linking ERP44 to ER stress adaptation. Dysregulation of ERP44 has been suggested to support tumor cell survival by enhancing ER redox balance and stress resistance [44].

Regulatory control by microRNAs further positions ERP44 within oncogenic miRNA-mRNA networks. In ACC, where ER stress and steroidogenic activity are prominent, aberrant ERP44 function may facilitate malignant progression by promoting protein quality control and survival signaling.

FAM32A (also known as OTAG-12 or ovarian tumor-associated gene 12) encodes a small nuclear protein that has been implicated in RNA metabolic pathways, including spliceosomal complexes [43, 44]. Functionally, FAM32A can induce G2 cell cycle arrest and apoptosis, thereby modulating sensitivity to chemotherapeutic agents [43, 44]. Dysregulation of FAM32A has been associated with tumor biology, where perturbations in pre-mRNA splicing and apoptotic regulation may contribute to oncogenesis. Endocrine-related phenotypic associations further support its potential relevance in endocrine malignancies. At the regulatory level, FAM32A is also subject to microRNA-mediated control, situating it within broader oncogenic regulatory networks. In ACC, abnormal FAM32A expression could influence cell cycle control and resistance to apoptosis, highlighting its potential role as an oncogenic biomarker and therapeutic target.

These four candidate oncogenes converge on key molecular axes directly relevant to adrenocortical carcinoma. CKS2 and FAM32A contribute to aberrant cell cycle control and apoptotic resistance, ERP44 integrates ER stress adaptation with proteostasis under the high steroidogenic load of adrenal cells, and ERG28 links sterol and steroid biosynthesis to endocrine metabolic regulation. Together, their dysregulation provides mechanistic plausibility for oncogenic transformation in ACC and supports their potential utility as endocrine-related biomarkers and therapeutic targets.

### 3.13 Pan-Cancer Expression Profiling of Candidate Genes

As part of a broader effort to understand the oncogenic relevance of our candidate genes across multiple cancer types, expression alterations were examined using TCGA data [20]. Approximate log fold-change (logFC) values were visually estimated from tumor versus normal expression boxplots available for 23 cancer types in TCGA [20]. This qualitative approach enabled large-scale expression shifts for CKS2, ERG28, FAM32A, and ERP44 to be captured, and insight was offered into whether these genes show consistent dysregulation beyond ACC. The summarized logFC profiles for all four genes across tumor types are shown in **Figure 5**.

**Figure 5.**
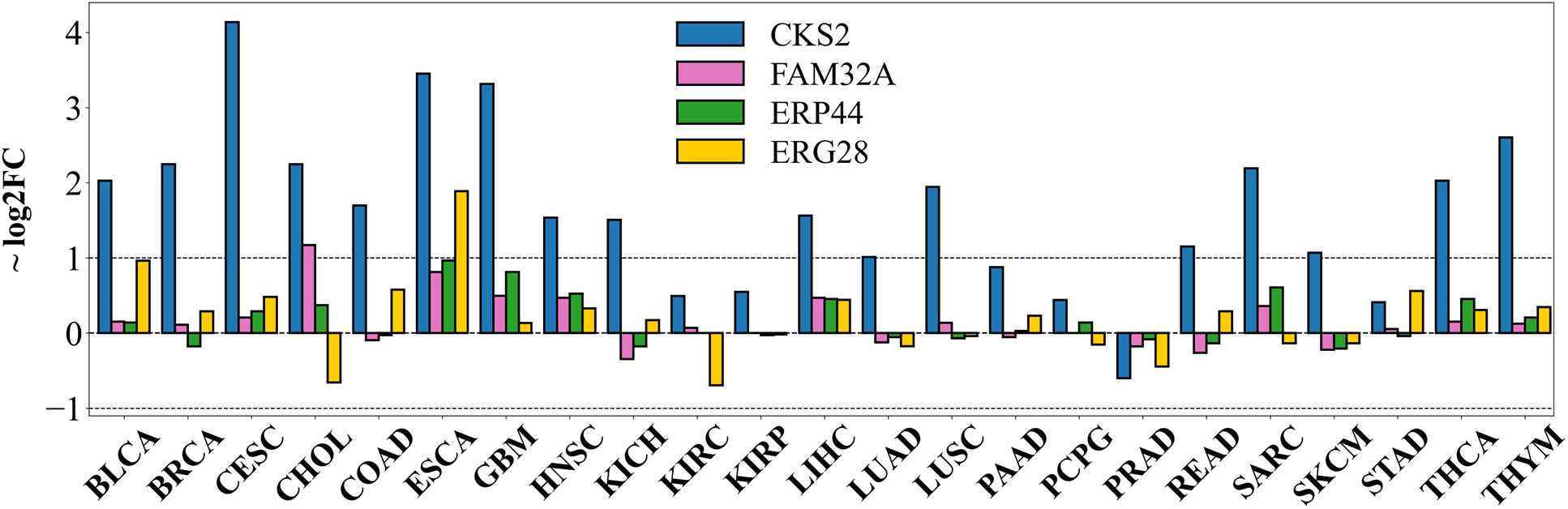
Pan-cancer log fold-change (logFC) expression of CKS2, ERG28, FAM32A, and ERP44 across 23 tumor types based on TCGA data [20].

Across TCGA cancers, CKS2 displayed striking and consistent upregulation, with particularly pronounced logFC values in cervical squamous cell carcinoma (CESC, 4.14), esophageal carcinoma (ESCA, 3.45), glioblastoma multiforme (GBM, 3.32), and thymoma (THYM, 2.60), as well as broad overexpression in multiple additional tumor types, thereby reinforcing its strong oncogenic profile beyond ACC. In contrast, FAM32A exhibited only subtle expression shifts, with modest gains observed in cholangiocarcinoma (CHOL, 1.17) and esophageal carcinoma (ESCA, 0.81), whereas alterations in most other cancers remained near-baseline, suggesting a context-dependent contribution to oncogenesis. ERP44 showed a pattern of moderate dysregulation, including elevated expression in ESCA (0.97), GBM (0.81), sarcoma (SARC, 0.61), and head and neck squamous cell carcinoma (HNSC, 0.52), supporting its potential role as an auxiliary oncogenic factor in select malignancies. ERG28 demonstrated a more heterogeneous landscape, with notable upregulation in ESCA (1.89), bladder cancer (BLCA, 0.96), colon adenocarcinoma (COAD, 0.58), and stomach adenocarcinoma (STAD, 0.56), but downregulation in kidney cancers (KIRC, -0.69; CHOL, -0.66; PRAD, -0.44), indicating that its dysregulation may reflect tissue-specific rather than universal oncogenic tendencies. These observations suggest that, while CKS2 emerges as a broadly oncogenic driver across multiple tumor contexts, the newly identified candidates FAM32A, ERP44, and ERG28 may represent ACC-specific oncogenic factors, warranting further mechanistic investigation and potential therapeutic exploration.

## 4. Discussion and Future Works

In this study, a novel systems-level framework was applied to elucidate post-transcriptional regulation in ACC, leading to the identification of four oncogene biomarkers (CKS2, ERG28, FAM32A, and ERP44) within tumor-specific ceRNA networks. The prioritization of CKS2 was supported not only by internal modeling but also by independent bioinformatic investigations that had previously characterized its elevated expression and poor prognostic association in ACC [48], whereas the other three candidate oncogenes have not been studied before. Prior studies in ACC have predominantly focused on classical cell cycle regulators, such as CCNB1, CDK1, TOP2A, and AURKA [49–51], yet the contribution of CKS2 has remained underexplored. While CKS2 is widely recognized for its role in cell cycle progression across diverse cancers [38, 42, 52], its functional relevance in ACC requires contextualization within the unique endocrine environment of the adrenal cortex. ACC cells are characterized by dysregulated steroid biosynthesis and altered endocrine signaling, both of which impose high metabolic and proliferative demands. CKS2 overexpression may facilitate these processes by sustaining aberrant CDK activity, thereby enabling rapid cell cycle transitions that accommodate the anabolic requirements of excessive steroidogenesis. This endocrine-specific framework provides a plausible mechanistic link between CKS2 dysregulation and the malignant phenotype of ACC.

Despite these insights, our study remains limited to computational and transcriptomic evidence. As correctly noted by the reviewer, functional assays are essential to establish causality. Gene knockdown or overexpression experiments in ACC cellular models, coupled with assessments of proliferation, apoptosis, and steroid output, will be required to verify whether CKS2 directly drives tumor progression through endocrine-specific mechanisms. Moreover, given the established interaction of CKS2 with cyclin-CDK complexes and checkpoint regulators [53, 54], evaluating the therapeutic relevance of CDK inhibitors or developing CKS2-targeted approaches in ACC could provide translational benefit.

Our findings introduce a ceRNA-based computational approach to biomarker discovery in ACC and propose CKS2 as a mechanistically plausible oncogenic driver whose role may be amplified by the unique metabolic and endocrine demands of adrenal cortical tumors. Future experimental validation will be crucial to confirm this hypothesis and to assess its therapeutic implications. Moreover, positioning candidate biomarkers such as CKS2 and ERP44 within the translational framework of companion diagnostics highlights their potential utility in precision oncology. As emphasized by McBrearty et al., robust biomarker validation and development of companion diagnostics are indispensable for accelerating drug development and ensuring patient stratification in clinical practice [6].

Among the four candidate oncogenes prioritized in this study, ERP44 (endoplasmic reticulum resident protein 44) represents a particularly compelling biomarker due to its localization within the ER lumen and its established role in protein folding, redox regulation, and unfolded protein response (UPR). In the context of ACC, which is an endocrine malignancy characterized by steroidogenesis, oxidative stress, and high secretory burden, aberrant expression of ERP44 may provide selective advantages to malignant cells. Indeed, our transcriptomic analyses revealed significant upregulation of ERP44 in ACC tumor tissues compared with normal adrenal cortex, with elevated expression strongly correlating with poor survival outcomes. The co-expression rewiring observed between ERP44, CKS2, and FAM32A in tumor samples further suggests that ERP44 is integrated into oncogenic transcriptional modules supporting malignant transformation.

Experimental evidence from other cancers provides mechanistic plausibility for this role. In gastric cancer, ERP44 overexpression was shown to suppress malignant phenotypes by promoting apoptosis and reducing proliferation through regulation of the eIF2α/CHOP stress axis [55]. Conversely, in oral squamous carcinoma and nasopharyngeal carcinoma, ERP44 was found to be pro-tumorigenic: its knockdown significantly inhibited proliferation, colony formation, and invasion, while in vivo experiments confirmed reduced xenograft growth [56, 57]. In breast and colorectal cancers, ERP44 upregulation has similarly been associated with stem-like properties and adverse prognosis [57]. These apparently divergent roles highlight the context-dependent function of ERP44, which may be shaped by the metabolic and secretory demands of different tumor types. In ACC, which is intrinsically reliant on ER capacity to sustain steroid hormone biosynthesis, ERP44 upregulation is likely co-opted to buffer ER stress, maintain proteostasis, and thereby promote tumor cell survival under the adverse microenvironmental conditions of rapid growth, hypoxia, and high metabolic turnover.

From a mechanistic perspective, the link between ERP44 and endocrine regulation is particularly noteworthy. Beyond its role in endocrine regulation, ERP44 has also been implicated in gliomas as an immunologically relevant oncogene, where its overexpression correlated with poor prognosis, altered immune cell infiltration, and checkpoint regulation [58]. This raises the possibility that in ACC, ERP44 may likewise modulate tumor-immune interactions in addition to buffering ER stress and supporting steroidogenic output. ERP44 participates in redox-dependent control of client proteins and has been reported to interact with IP3R1 to modulate calcium flux [55]. Given that steroid hormone synthesis in the adrenal cortex is tightly regulated by calcium-dependent signaling and ER chaperone activity, dysregulated ERP44 may directly contribute to altered steroidogenic output in ACC. This provides a rational integration of ERP44 function with the pathological hallmarks of ACC beyond general oncogenic proliferation, addressing the reviewer’s concern regarding cancer-type specificity.

Nonetheless, the absence of direct functional assays in ACC remains a limitation of this study. Although our computational and clinical evidence robustly implicates ERP44 as an oncogenic driver, causal inference requires experimental validation. Specifically, knockdown or overexpression experiments in ACC cell models, coupled with assays of proliferation, apoptosis, steroid synthesis, and ER stress markers, will be critical to establish whether ERP44 directly mediates the malignant phenotype of ACC. In vivo studies using xenograft or patient-derived models could further clarify its contribution to tumor progression and therapy resistance.

In summary, ERP44 emerges as a novel ACC-associated biomarker whose elevated expression correlates with poor prognosis and whose functional profile is well aligned with the ER- and steroid-centric biology of the adrenal cortex. In support of its translational relevance, exosomal ERP44 derived from ER-stressed cells has been shown to strengthen cisplatin resistance in nasopharyngeal carcinoma [59]. Given the inherently secretory nature of ACC, a similar exosome-mediated mechanism may contribute to therapeutic resistance in adrenal tumors, underscoring the clinical value of targeting ERP44. While supportive evidence from other tumor systems suggests that ERP44 can regulate proliferation, invasion, apoptosis, and stress adaptation, future ACC-specific functional studies are required to confirm causality and to evaluate the translational potential of ERP44 as a therapeutic target. This need for functional validation is consistent with recent bioinformatic investigations in other cancer contexts, such as the study by Duan et al. on PIEZO1 in non-small cell lung cancer, where in silico predictions of regulatory networks were proposed but required experimental confirmation to establish causality [12].

FAM32A was identified as significantly upregulated in ACC tumors and strongly associated with poor prognosis in our cohort. Functional annotations linked FAM32A to RNA binding and apoptotic regulation, consistent with its putative role in cell fate determination. In gastric cancer, suppression of FAM32A reduced apoptosis and induced chemoresistance by attenuating the p53 signaling pathway [60]. Within the endocrine context of ACC, such dysregulation of apoptosis could facilitate tumor cell survival under conditions of endoplasmic reticulum stress and steroidogenic metabolic load, processes that normally trigger programmed cell death in adrenal cortical cells. The tumor-specific co-regulation observed between FAM32A and CKS2 in our study further suggests that FAM32A may integrate into proliferative signaling circuits, promoting malignant progression. However, despite these associations, no experimental validation has yet confirmed its causal role in ACC, emphasizing the need for functional assays such as gene knockdown or overexpression in adrenal tumor models.

ERG28 encodes a conserved endoplasmic reticulum membrane protein essential for sterol biosynthesis, and its strong evolutionary conservation from yeast to humans underscores its fundamental role in lipid metabolism [61]. Our results demonstrated robust overexpression of ERG28 in ACC and its significant association with poor survival outcomes. This is particularly relevant to ACC, where cholesterol metabolism is the cornerstone of steroid hormone synthesis. Aberrant ERG28 activity may enhance the flux of sterol intermediates, thereby fueling both membrane biogenesis and hormone overproduction, two defining pathological features of ACC. The coupling of metabolic rewiring with accelerated proliferation provides a plausible mechanistic explanation for its oncogenic behavior in this malignancy. While studies in yeast have demonstrated severe growth defects upon ERG28 disruption [61], functional studies in adrenal cancer models remain absent. Future investigations employing metabolic flux analysis and ERG28 silencing could determine whether it serves as a driver of both tumor aggressiveness and endocrine dysregulation in ACC.

FAM32A and ERG28 underscore how candidate oncogenes in ACC extend beyond generic cell cycle regulators and converge on endocrine-specific vulnerabilities: FAM32A aligns with apoptotic escape under stress, while ERG28 directly links to steroid/sterol metabolism, a hallmark of adrenal physiology and tumor pathology. By situating these candidates within the unique metabolic and endocrine framework of ACC, our findings provide mechanistic plausibility to their oncogenic potential. Nevertheless, consistent with the reviewer’s observation, the absence of cellular functional assays remains a limitation, and future experimental work will be essential to confirm causality and therapeutic relevance.

## 5. Conclusion

In this study, an integrative systems biology framework was employed to unravel post-transcriptional regulatory mechanisms in ACC. Through ceRNA network modeling, transcriptomic validation, survival analyses, and functional annotation, four candidate oncogenes; CKS2, FAM32A, ERP44, and ERG28 were prioritized. All four genes were found to be significantly upregulated in tumor tissues compared with normal adrenal samples, to exhibit tumor-specific co-expression modules, and to be associated with markedly reduced overall survival. Their enrichment in pathways governing cell cycle progression, apoptosis regulation, proteostasis, and sterol biosynthesis underscores their biological relevance in malignant transformation. These findings highlight CKS2, FAM32A, ERP44, and ERG28 as promising diagnostic and prognostic biomarkers with potential therapeutic implications in ACC.

Despite these advances, several limitations must be acknowledged. The analyses were constrained by the limited sample size of TCGA-ACC, which may reduce statistical power and generalizability. Furthermore, the absence of in vitro or in vivo functional validation precludes definitive causal inferences regarding the oncogenic roles of these candidates. Potential confounding effects related to patient heterogeneity, including racial and demographic differences, were not explicitly addressed and warrant consideration in future studies. Therefore, while the present findings provide a mechanistic framework and nominate robust biomarker candidates, experimental validation and larger, demographically diverse cohorts will be essential to fully establish their translational value in ACC.

## Data Availability

All data produced in the present work are contained in the manuscript.

## Funding Statement

No specific grant was received for this research.

## Author Statement

J. O.: Conceptualization, Data curation, Formal analysis, Investigation, Methodology, Project administration, Resources, Software, Supervision, Validation, Visualization, Writing - original draft, Writing - review and editing

## Ethics Statement

N/A

## Conflict of Interest Statement

The author declares no conflicts of interest.

## Notes

### Competing Interest Statement

The authors have declared no competing interest.

### Funding Statement

This study did not receive any funding.

### Author Declarations

This study used only publicly available, de-identified human transcriptomic data that were openly accessible prior to the initiation of the study. Adrenocortical carcinoma (ACC) data were obtained from the open-access portion of The Cancer Genome Atlas (TCGA) data portal. Normal adrenal gland transcriptomic data were obtained from the publicly released Genotype-Tissue Expression (GTEx) project. All datasets were accessed without any request for controlled access, IRB approval, or additional authorization. No new data were generated, and no direct involvement of human participants occurred in this study.

### Summary of Updates

This revised version of the manuscript incorporates several updates to improve clarity alignment and overall coherence with the core scientific message of the study. First the manuscript title has been revised to more accurately reflect the content scope and conceptual foundation of the work ensuring closer alignment between the title and the underlying analyses and conclusions presented in the paper. Second additional recent publications from the authors research program have been incorporated into the reference list. These studies provide important contextual background and methodological continuity and are expected to facilitate a clearer and deeper understanding of the current work for readers. Third the table summarizing all functional annotations has been removed. The most relevant and biologically meaningful annotations are now explicitly discussed within the main text while inclusion of the complete annotation set was found to be redundant potentially confusing and in some cases not directly related to the central questions addressed in the manuscript. This change was made to improve focus readability and conceptual clarity while preserving all key scientific insights.

## References

[1] Allolio, Bruno, and Martin Fassnacht. “Adrenocortical carcinoma: clinical update.” The Journal of Clinical Endocrinology & Metabolism 91, no. 6 (2006): 2027–2037.

[2] Kerkhofs, Thomas MA, Rob HA Verhoeven, Jan Maarten Van der Zwan, Jeanne Dieleman, Michiel N. Kerstens, Thera P. Links, Lonneke V. Van de Poll-Franse, and Harm R. Haak. “Adrenocortical carcinoma: a population-based study on incidence and survival in the Netherlands since 1993.” European Journal of Cancer 49, no. 11 (2013): 2579–2586.

[3] Terzolo, Massimo, Alberto Angeli, Martin Fassnacht, Fulvia Daffara, Libuse Tauchmanova, Pier Antonio Conton, Ruth Rossetto et al. “Adjuvant mitotane treatment for adrenocortical carcinoma.” New England Journal of Medicine 356, no. 23 (2007): 2372–2380.

[4] Fassnacht, Martin, Rossella Libé, Matthias Kroiss, and Bruno Allolio. “Adrenocortical carcinoma: a clinician’s update.” Nature Reviews Endocrinology 7, no. 6 (2011): 323–335.

[5] Decmann, Abel, Pál Perge, Peter Istvan Turai, Attila Patócs, and Peter Igaz. “Non-coding RNAs in adrenocortical cancer: from pathogenesis to diagnosis.” Cancers 12, no. 2 (2020): 461.

[6] McBrearty, Noreen, Devika Bahal, and Suso Platero. “Fast-tracking drug development with biomarkers and companion diagnostics.” Journal of Cancer Metastasis and Treatment 10 (2024): N-A.

[7] Slack, Frank J., and Arul M. Chinnaiyan. “The role of non-coding RNAs in oncology.” Cell 179, no. 5 (2019): 1033–1055.

[8] Salimian, Niloufar, Maryam Peymani, Kamran Ghaedi, Sepideh Mirzaei, and Mehrdad Hashemi. “Diagnostic and therapeutic potential of LINC01929 as an oncogenic LncRNA in human cancers.” Pathology-Research and Practice 244 (2023): 154409.

[9] Sayaf, Hossein, Niloufar Salimian, Mahnaz Mohammadi, Parisa Ahmadi, Amir Gholamzad, Sadegh Babashah, Maliheh Entezari, Najma Farahani, Maryam Montazeri, and Mehrdad Hashemi. “Botox- A induced apoptosis and suppressed cell proliferation in fibroblasts pre-treated with breast cancer exosomes.” Molecular and Cellular Probes 79 (2025): 102007.

[10] Tay, Yvonne, John Rinn, and Pier Paolo Pandolfi. “The multilayered complexity of ceRNA crosstalk and competition.” Nature 505, no. 7483 (2014): 344–352.

[11] Yang, Ni, Kuo Liu, Mengxuan Yang, and Xiang Gao. “ceRNAs in cancer: mechanism and functions in a comprehensive regulatory network.” Journal of oncology 2021, no. 1 (2021): 4279039.

[12] Duan, Lingdi, Min Zhao, Hongquan Wei, Wei Dong, Xiaomin Bi, Lin Ang, and Shan Zhang. “Bioinformatics analysis of the association between miR-942-5p-induced downregulation of PIEZO-type mechanosensitive ion channel component 1 and poor prognosis in non-small cell lung cancer mediated by the mitogen-activated protein kinase pathway signaling pathway.” Oncology and Translational Medicine 10, no. 6 (2024): 272–280.

[13] Vidal, Marc, Michael E. Cusick, and Albert-László Barabási. “Interactome networks and human disease.” Cell 144, no. 6 (2011): 986–998.

[14] Qi, Xin, Yuxin Lin, Jiajia Chen, and Bairong Shen. “Decoding competing endogenous RNA networks for cancer biomarker discovery.” Briefings in Bioinformatics 21, no. 2 (2020): 441–457.

[15] Li, Xing, Bing Li, Pixin Ran, and Lanying Wang. “Identification of ceRNA network based on a RNA-seq shows prognostic lncRNA biomarkers in human lung adenocarcinoma.” Oncology Letters 16, no. 5 (2018): 5697–5708.

[16] Omidi, Javad. “Synergistic dual oncogenic role of CKS2 and ACAT2: Enhanced regulatory coherence and biomarker potential in adrenocortical carcinoma.” Cancer Genetics (2025).

[17] Omidi, Javad. “miR-507 and miR-665 as central MicroRNA regulators in the ceRNA network of adrenocortical carcinoma: A systems biology approach.” Human Gene (2025): 201498.

[18] Omidi, Javad. “miR-466 as a Central miRNA with Tumor-Suppressive Potential in the Regulatory Network of Adrenocortical Carcinoma.” In Silico Research in Biomedicine (2026): 100214.

[19] Omidi, Javad. “Molecular Landscape and Biomarker Discovery in Adrenocortical Carcinoma: An Integrative Review of Bioinformatics and Translational Insights.” Pathology-Research and Practice (2025): 156295.

[20] The Cancer Genome Atlas (TCGA). 2024. “TCGA Program.” National Cancer Institute. https://www.cancer.gov/about-nci/organization/ccg/research/structural-genomics/tcga

[21] Assié, Guillaume, Eric Letouzé, Martin Fassnacht, Anne Jouinot, Windy Luscap, Olivia Barreau, Hanin Omeiri et al. “Integrated genomic characterization of adrenocortical carcinoma.” Nature genetics 46, no. 6 (2014): 607–612.

[22] Feige, J., and N. Cherradi. “Serum miR-483-5p and miR-195 are predictive of recurrence risk in adrenocortical cancer patients.” Endocr. Relat. Cancer 20 (2013): 579–594.

[23] Özata, Deniz M., Stefano Caramuta, David Velazquez-Fernandez, Pinar Akçakaya, Hong Xie, Anders Höög, Jan Zedenius, Martin Bäckdahl, Catharina Larsson, and Weng-Onn Lui. “The role of microRNA deregulation in the pathogenesis of adrenocortical carcinoma.” Endocrine-related cancer 18, no. 6 (2011): 643–655.

[24] GTEx 2025 Consortium, Kristin G. Ardlie, David S. Deluca, Ayellet V. Segrè, Timothy J. Sullivan, Taylor R. Young, Ellen T. Gelfand et al. “The Genotype-Tissue Expression (GTEx 2025) pilot analysis: multitissue gene regulation in humans.” Science 348, no. 6235 (2015): 648–660.

[25] Ritchie, Matthew E., Belinda Phipson, D. I. Wu, Yifang Hu, Charity W. Law, Wei Shi, and Gordon K. Smyth. “limma powers differential expression analyses for RNA-sequencing and microarray studies.” Nucleic acids research 43, no. 7 (2015): e47–e47.

[26] Sha, Ying, John H. Phan, and May D. Wang. “Effect of low-expression gene filtering on detection of differentially expressed genes in RNA-seq data.” In 2015 37th Annual International Conference of the IEEE Engineering in Medicine and Biology Society (EMBC), pp. 6461–6464. IEEE, 2015.

[27] Conesa, Ana, Pedro Madrigal, Sonia Tarazona, David Gomez-Cabrero, Alejandra Cervera, Andrew McPherson, Michał Wojciech Szcześniak et al. “A survey of best practices for RNA-seq data analysis.” Genome biology 17, no. 1 (2016): 13.

[28] Ru, Yuanbin, Katerina J. Kechris, Boris Tabakoff, Paula Hoffman, Richard A. Radcliffe, Russell Bowler, Spencer Mahaffey et al. “The multiMiR R package and database: integration of microRNA-target interactions along with their disease and drug associations.” Nucleic acids research 42, no. 17 (2014): e133–e133.

[29] Tong, Feng, Youhua Ying, Haihua Pan, Wei Zhao, Hongchen Li, and Xiaoli Zhan. “MicroRNA-466 (miR-466) functions as a tumor suppressor and prognostic factor in colorectal cancer (CRC).” Bosnian journal of basic medical sciences 18, no. 3 (2018): 252.

[30] Colden, Melissa, Altaf A. Dar, Sharanjot Saini, Priya V. Dahiya, Varahram Shahryari, Soichiro Yamamura, Yuichiro Tanaka, Gary Stein, Rajvir Dahiya, and Shahana Majid. “MicroRNA-466 inhibits tumor growth and bone metastasis in prostate cancer by direct regulation of osteogenic transcription factor RUNX2.” Cell death & disease 8, no. 1 (2018): e2572–e2572.

[31] Li, Hongxiang, Yin Li, Dongmei Tian, Jiaqian Zhang, and Shiwei Duan. “miR-940 is a new biomarker with tumor diagnostic and prognostic value.” Molecular Therapy Nucleic Acids 25 (2021): 53–66.

[32] Fang, Xin, and Anping Pan. “MiR-507 inhibits the progression of gastric carcinoma via targeting CBX4-mediated activation of Wnt/β-catenin and HIF-1α pathways.” Clinical and Translational Oncology 24, no. 10 (2022): 2021–2028.

[33] Huang, Hsi-Yuan, Yang-Chi-Dung Lin, Jing Li, Kai-Yao Huang, Sirjana Shrestha, Hsiao-Chin Hong, Yun Tang, et al. “miRTarBase 2020: updates to the experimentally validated microRNA-target interaction database.” Nucleic acids research 48, no. D1 (2020): D148-D154.

[34] Chen, Yuhao, and Xiaowei Wang. “miRDB: an online database for prediction of functional microRNA targets.” Nucleic acids research 48, no. D1 (2020): D127–D131.

[35] Agarwal, Vikram, George W. Bell, Jin-Wu Nam, and David P. Bartel. “Predicting effective microRNA target sites in mammalian mRNAs.” elife 4 (2015): e05005.

[36] Liu, Cheng, Chun Yu, Guoxin Song, Xingchen Fan, Shuang Peng, Shiyu Zhang, Xin Zhou et al. “Comprehensive analysis of miRNA-mRNA regulatory pairs associated with colorectal cancer and the role in tumor immunity.” BMC genomics 24, no. 1 (2023): 724.

[37] Szklarczyk, Damian, Andrea Franceschini, Stefan Wyder, Kristoffer Forslund, Davide Heller, Jaime Huerta-Cepas, Milan Simonovic et al. “STRING v10: protein-protein interaction networks, integrated over the tree of life.” Nucleic acids research 43, no. D1 (2015): D447–D452.

[38] Lai, Yueliang, and Ye Lin. “Biological functions and therapeutic potential of CKS2 in human cancer.” Frontiers in Oncology 14 (2024): 1424569.

[39] D’Amore, Claudio, Christian Borgo, Stefania Sarno, and Mauro Salvi. “Role of CK2 inhibitor CX-4945 in anti-cancer combination therapy-potential clinical relevance.” Cellular Oncology 43 (2020): 1003–1016.

[40] Marschke, R. F., M. J. Borad, R. W. McFarland, R. H. Alvarez, J. K. Lim, C. S. Padgett, D. D. Von Hoff, S. E. O’Brien, and D. W. Northfelt. “Findings from the phase I clinical trials of CX-4945, an orally available inhibitor of CK2.” Journal of Clinical Oncology 29, no. 15_suppl (2011): 3087–3087.

[41] Borgo, Christian, Claudio D’Amore, Stefania Sarno, Mauro Salvi, and Maria Ruzzene. “Protein kinase CK2: a potential therapeutic target for diverse human diseases.” Signal transduction and targeted therapy 6, no. 1 (2021): 183.

[42] Qiu, Danqi, Hongshi Cai, Jianfeng Liang, Ziyi Wang, Fan Song, Yaoqi Jiang, Rukeng Tan, and Jingsong Hou. “Identification of CKS2 as a novel prognostic biomarker and potential therapeutic target for oral squamous cell carcinoma.” Translational Cancer Research 12, no. 9 (2023): 2276.

[43] “UniProt: the Universal protein knowledgebase in 2025.” Nucleic Acids Research 53, no. D1 (2025): D609–D617.

[44] Safran, Marilyn, Irina Dalah, Justin Alexander, Naomi Rosen, Tsippi Iny Stein, Michael Shmoish, Noam Nativ et al. “GeneCards Version 3: the human gene integrator.” Database 2010 (2010).

[45] Anelli, Tiziana, Massimo Alessio, Alexandre Mezghrani, Thomas Simmen, Fabio Talamo, Angela Bachi, and Roberto Sitia. “ERp44, a novel endoplasmic reticulum folding assistant of the thioredoxin family.” The EMBO journal (2002).

[46] Zhang, Jianchao, Qinyu Zhu, Xi’E. Wang, Jiaojiao Yu, Xinxin Chen, Jifeng Wang, Xi Wang, Junyu Xiao, Chih-chen Wang, and Lei Wang. “Secretory kinase Fam20C tunes endoplasmic reticulum redox state via phosphorylation of Ero1α.” The EMBO Journal 37, no. 14 (2018): e98699.

[47] Higo, Takayasu, Mitsuharu Hattori, Takeshi Nakamura, Tohru Natsume, Takayuki Michikawa, and Katsuhiko Mikoshiba. “Subtype-specific and ER lumenal environment-dependent regulation of inositol 1, 4, 5-trisphosphate receptor type 1 by ERp44.” Cell 120, no. 1 (2005): 85–98.

[48] Yin, Mengsha, Yao Wang, Xinhua Ren, Mingyue Han, Shanshan Li, Ruishuang Liang, Guixia Wang, and Xiaokun Gang. “Identification of key genes and pathways in adrenocortical carcinoma: evidence from bioinformatic analysis.” Frontiers in Endocrinology 14 (2023): 1250033.

[49] Li, Xiunan, Jiayi Li, Leizuo Zhao, Zicheng Wang, Peizhi Zhang, Yingkun Xu, and Guangzhen Wu. “Comprehensive Multiomics Analysis Reveals Potential Diagnostic and Prognostic Biomarkers in Adrenal Cortical Carcinoma.” Computational and Mathematical Methods in Medicine 2022, no. 1 (2022): 2465598.

[50] Guo, Jinshuai, Yinzhong Gu, Xiaoyu Ma, Lu Zhang, Huimin Li, Zhongyi Yan, Yali Han, Longxiang Xie, and Xiangqian Guo. “Identification of hub genes and pathways in adrenocortical carcinoma by integrated bioinformatic analysis.” Journal of cellular and molecular medicine 24, no. 8 (2020): 4428–4438.

[51] Xing, Zengmiao, Zuojie Luo, Haiyan Yang, Zhenxing Huang, and Xinghuan Liang. “Screening and identification of key biomarkers in adrenocortical carcinoma based on bioinformatics analysis.” Oncology letters 18, no. 5 (2019): 4667–4676.

[52] Yu, Min-Hao, Yang Luo, Shao-Lan Qin, Zheng-Shi Wang, Yi-Fei Mu, and Ming Zhong. “Up-regulated CKS2 promotes tumor progression and predicts a poor prognosis in human colorectal cancer.” American journal of cancer research 5, no. 9 (2015): 2708.

[53] You, Hanyu, Huayue Lin, and Zhongying Zhang. “CKS2 in human cancers: Clinical roles and current perspectives.” Molecular and clinical oncology 3, no. 3 (2015): 459–463.

[54] Xu, J-H., Y. Wang, and D. Xu. “CKS2 promotes tumor progression and metastasis and is an independent predictor of poor prognosis in epithelial ovarian cancer.” European Review for Medical & Pharmacological Sciences 23, no. 8 (2019).

[55] Tian, Yongjing, Haibin Sun, Yinshengboer Bao, Haiping Feng, Jian Pang, Riletu En, Hongliang Jiang, and Tengqi Wang. “ERp44 regulates the proliferation, migration, invasion, and apoptosis of gastric cancer cells via activation of ER stress.” Biochemical Genetics 61, no. 2 (2023): 809–822.

[56] Cho, Jin Hyoung, Young-Joo Jeon, Seon-Min Park, Jae-Cheon Shin, Tae-Hoon Lee, Seunggon Jung, Hongju Park et al. “Multifunctional effects of honokiol as an anti-inflammatory and anti-cancer drug in human oral squamous cancer cells and xenograft.” Biomaterials 53 (2015): 274–284.

[57] Tian, Hui, Si Shi, Bo You, Qicheng Zhang, Miao Gu, and Yiwen You. “ER resident protein 44 promotes malignant phenotype in nasopharyngeal carcinoma through the interaction with ATP citrate lyase.” Journal of Translational Medicine 19, no. 1 (2021): 77.

[58] Ji, Xiang, Zhenglou Chen, Yunjiang Wang, Xuqi Huo, Xiaodong Liang, Hongsheng Wang, and Min Xu. “ERP44 could serve as a bridge mediating prognosis and immunity for glioma via single-cell and bulk RNA-sequencing.” Gene 933 (2025): 148963.

[59] Xia, Tian, Hui Tian, Kaiwen Zhang, Siyu Zhang, Wenhui Chen, Si Shi, and Yiwen You. “Exosomal ERp44 derived from ER-stressed cells strengthens cisplatin resistance of nasopharyngeal carcinoma.” BMC cancer 21, no. 1 (2021): 1003.

[60] Agatsuma, Yuya, Dai Shimizu, Shinichi Umeda, Haruyoshi Tanaka, Norifumi Hattori, Masamichi Hayashi, Mitsuro Kanda et al. “FAM32A Suppression Decreases 5-Fluorouracil-induced Apoptosis and Is Associated With Poor Prognosis in Gastric Cancer.” Cancer Genomics & Proteomics 22, no. 1 (2025): 55–69.

[61] Ottolenghi, Chris, Iraj Daizadeh, Albert Ju, Sophia Kossida, Georges Renault, Michel Jacquet, Arlette Fellous, Walter Gilbert, and Reiner Veitia. “The genomic structure of C14orf1 is conserved across eukarya.” Mammalian Genome 11, no. 9 (2000): 786–788.3

